# Evaluation of ultrasound as diagnostic tool in patients with clinical features suggestive of carpal tunnel syndrome in comparison to nerve conduction studies: study protocol for a diagnostic testing study

**DOI:** 10.1101/2023.01.19.23284770

**Authors:** María de la Paz Murciano Casas, Manuel Rodríguez-Piñero, Aguas-Santas Jiménez Sarmiento, Mercedes Álvarez López, Gema Jiménez Jurado

## Abstract

**Background:** Carpal Tunnel Syndrome (CTS) is the most common compressive neuropathy, accounting for 90% of all neuropathies. Its prevalence ranges from 3.8% - 7.8% in the population. The gold standard for its diagnosis is the neurophysiological study (85% sensitivity and 95% specificity), with the disadvantage of being invasive, complex and expensive, which means an increase in cost and time for the diagnosis of the disease. The main objective of this diagnostic test evaluation study is to investigate the value of ultrasound in the diagnosis of CTS, and among the secondary objectives, to establish the ultrasound parameters that are predictors of CTS in comparison with neurophysiological studies, attempting to standardize a protocol and reference values that determine the presence or absence of CTS.

**Methods:** Prospective, cross-sectional study. The reference test with which we compared the ultrasound is the neurophysiological test (NPT). Patients will come consecutively from the Neurophysiology Department of the Virgen Macarena Hospital, with clinical suspicion of CTS and fulfilling the inclusion/exclusion criteria. To calculate the sample size (EPIDAT program) we proposed a sensitivity of 78% and specificity of 87% with a confidence level of 95%, requiring 438 patients (264 NPT positive, 174 NPT negative). We followed an ultrasound study protocol that included the ultrasound variables: cross-sectional area at the entrance and exit of the tunnel, range of nerve thinning, wrist-forearm index, flexor retinaculum bulging, power Doppler uptake and the existence of adjacent wrists or masses. We propose a timeline for the study to be performed between 2020 and 2023. Finally, we propose a cost-effectiveness analysis.

**Discussion:** Ultrasound not only allows to objectify the alterations of the median nerve but also the underlying pathological mechanisms in CTS. A multitude of ultrasound parameters have been described that should be regarded in syndrome’s study, among which we included the cross-sectional area, the range of nerve thinning, the wrist-forearm index, flexor retinaculum bulging, power Doppler uptake and assessment of anatomical alterations. The use of ultrasound as a diagnostic tool in CTS has many advantages for both doctors and the patients, as it is a non-invasive, convenient, and fast tool increasingly accessible to professionals.

**Administrative information:** 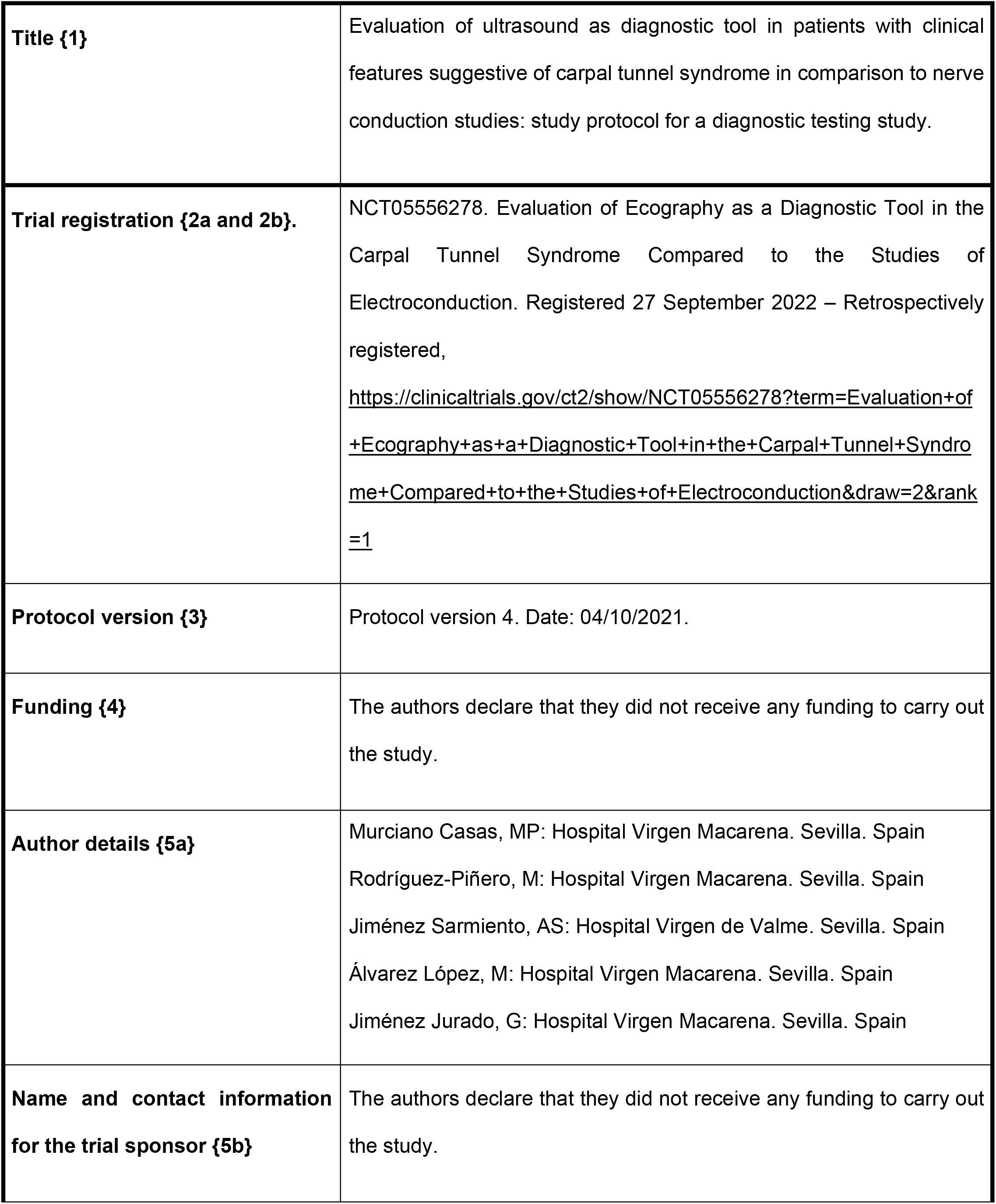
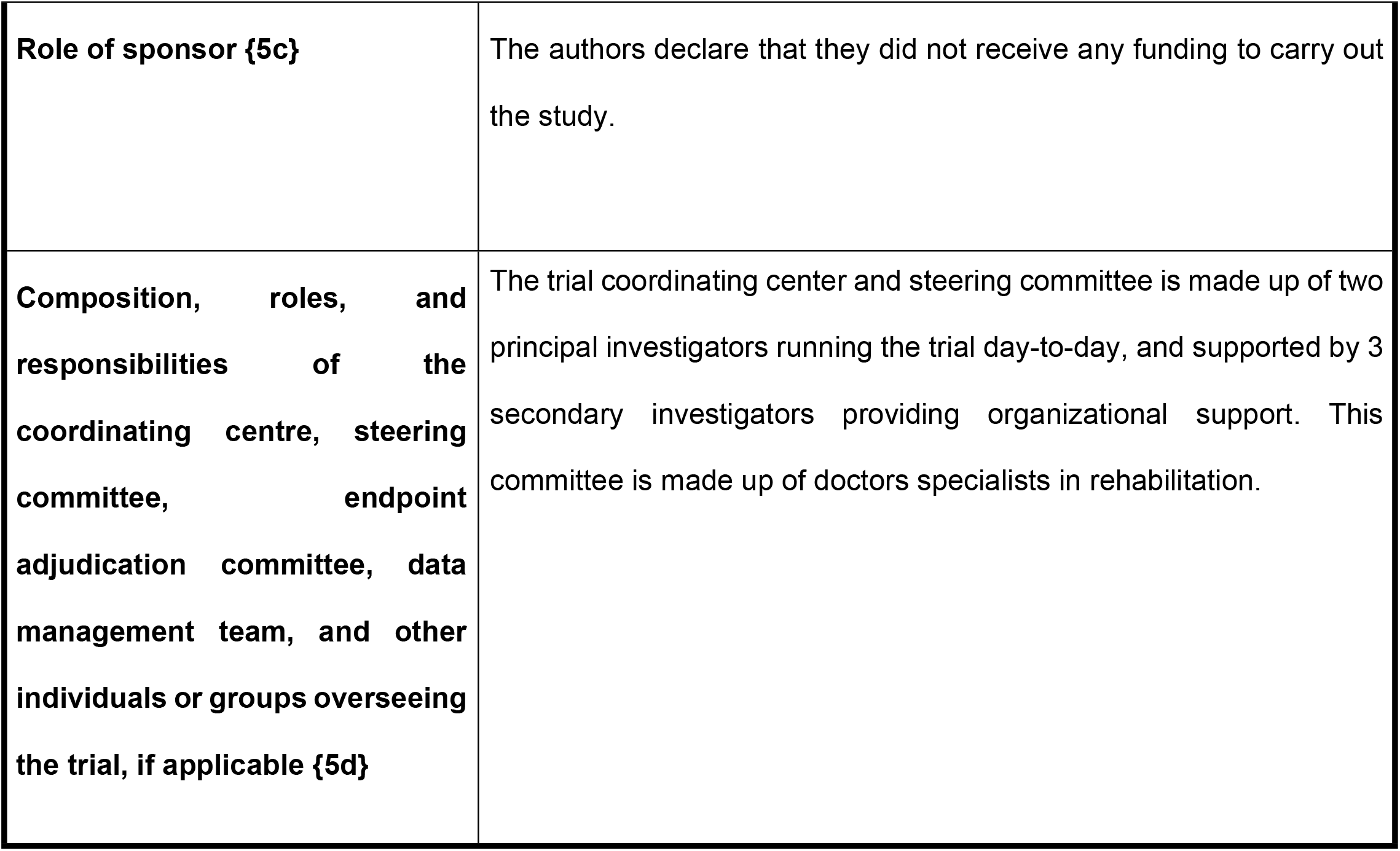

## Introduction

### Background and rationale {6a}

The carpal tunnel is an anatomical space located at the base of the palm of the hand, delimited by a rigid floor and walls, the carpal bones, and a roof with a certain capacity for distension, the transverse carpal ligament or flexor retinaculum. The entrance of the tunnel is located between the scaphoid and pyramidal bones, and its exit between the trapezium and hamate bones. A number of anatomical structures pass through this relatively narrow space as it transitions from the forearm to the hand: the tendons of the superficial and deep flexor muscles of the fingers, the tendon of the long flexor muscle of the first finger, and the median nerve (1). Thus, an increase in volume within this space, however small, can lead to compression of the nerve (2).

The dimensions of the carpal tunnel are 20 mm at the level of the hook of the hamate, narrow compared to the proximal (24 mm) or distal (25 mm) dimensions (4). In healthy individuals the intratunnel pressure ranges between 3-5 mmHg in neutral position (5); vascular compromise of the nerve begins when the pressure exceeds 20-30 mmHg (6), which could be favored by certain postures or gestures as common as using a computer mouse (7). Adjacent musculature may also play a role in the etiology; in a cadaver study, the muscle belly of the flexor digitorum superficialis was observed to access the proximal tunnel during wrist extension, similar to the lumbrical muscles observed in contact with the distal tunnel during metacarpophalangeal flexion (8).

The median nerve is responsible for the sensory innervation of the first three fingers and the radial border of the fourth, which is why patients who suffer compression of the median nerve inside the tunnel report pain and hypoesthesia in these fingers. The palm of the hand, however, usually remains unaltered, as it is innervated by a sensory cutaneous branch that emerges 6 cm proximal to the entrance of the nerve into the tunnel (5). Carpal Tunnel Syndrome (CTS) is the most common compressive neuropathy, accounting for 90% of all neuropathies (1,9). It is estimated that 1 in 5 patients with symptoms of pain, paresthesia and numbness of the hand will be diagnosed with CTS based on examination and an electrodiagnostic test (9).

Its prevalence ranges from 3.8% to 7.8% of the general population, being more prevalent in females than in males (10% vs. 5.8%), with an incidence of 9.3 new cases per 100 persons per year (10,11). It leads to increased absenteeism, morbidity and, depending on the country, a cost of up to 2 billion dollars per year (11). It is characterized by a combination of motor and sensory symptomatology leading to functional impairment, manifested by weakness of the intrinsic hand musculature with decreased grip strength, pain and paresthesia (5). The etiology of this entity remains uncertain, and most cases are labelled idiopathic; recently, magnetic resonance imaging, biomechanical and histological studies have proposed a close relationship between neuronal vascular dysfunction, synovial tissue, flexor tendons and their sheaths, with the onset of the syndrome (3).

Diagnosis has traditionally been established by anamnesis and physical examination; electrodiagnostic testing is frequently used as a method of confirmation and assessment of severity. Although these studies offer 85% sensitivity and 95% specificity (12), they have the disadvantage of being invasive and unpleasant for patients. Recent advances in ultrasonography techniques have considerably improved the quality of ultrasound imaging of the nerve while providing more compact and economical equipment that makes it more accessible to the clinician, making ultrasonography a potential tool for the evaluation of entrapment neuropathies.

The high prevalence of CTS in the general population causes a high demand for neurophysiological studies, which to date is considered the gold standard in its diagnosis; this leads to long waiting times for the test to be carried out as well as an increase in direct and indirect costs derived from the disease. The availability of an additional technique that allows CTS to be diagnosed quickly, non-invasively and inexpensively would represent a clear advance in healthcare and an improvement in the efficiency of the healthcare system. In this sense, the present study aims to investigate the diagnostic value of ultrasound in the diagnosis of CTS, the specific ultrasound parameters that predict the existence of the syndrome in comparison with neurophysiological studies, and thus try to standardize a study protocol and reference values that determine the presence or absence of the syndrome.

### Explanation for the choice of comparators {6b}

We compared the NPT based on the recommendations of the American Association of Electrodiagnostic Medicine (AAEM) protocol with the ultrasound study protocol described in Chen et al. (11) for assessing its effectiveness. The study protocol included different variables: cross-sectional area at the entrance and exit of the tunnel, range of nerve thinning, wrist-forearm index, flexor retinaculum bulging, power Doppler uptake and the existence of adjacent wrists or masses.

## Objectives {7}

### Primary

To assess the efficacy of ultrasound in the diagnosis of CTS with respect to nerve conduction studies.

### Secondary

- To determine the set of ultrasound parameters (ultrasound standards and reference values) that best predict the existence of CTS.
- To correlate the intensity or degree of involvement between ultrasound findings and nerve conduction studies.
- To propose a standardised protocol for the study of CTS by ultrasound.
- To establish the interobserver and intraobserver variability of ultrasound in the diagnosis of CTS.
- To perform a cost-effectiveness analysis between ultrasound and neurophysiological studies in the diagnosis of CTS.
- To detect and describe anatomical alterations and pathological findings that could predispose to the presence of CTS.

#### Hypothesis

Ultrasound is more effective than neurophysiological studies in the diagnosis of CTS.

### Trial design {8}

Prospective, cross-sectional study that compares two diagnostic tests: electroneurography and ultrasound in the diagnosis of CTS. Also, a cost-effectiveness analysis will be proposed to evaluate the costs of the consumables established by the Andalusian Health Service used in both tests and the average time for each of the tests, in order to calculate the cost of the professional when performing each of them.

## Methods: Participants, interventions and outcomes

### Study setting {9}

Patients will come consecutively from the Neurophysiology Department of the Virgen Macarena Hospital in Seville (Spain), with clinical suspicion of CTS and fulfilling the inclusion/exclusion criteria. Electroneurography or ultrasound diagnosis techniques will be performed to compare their effectiveness.

### Eligibility criteria {10}

We establish the following inclusion criteria in order to be able to take part in the study:

1. Presenting clinical symptoms suggestive of CTS, at least one of the following:

- Neuropathic profile pain in wrist-hand at the level of the median nerve territory (wrist, first-second-third, and radial half of the fourth finger by palmar and/or volar side), possibility of irradiation of the pain towards the forearm and/or arm.
- Nocturnal symptoms.
- Hypotrophy or atrophy of the thenar eminence.
- Self-described sensory alteration (sensation of cramping, numbness, tingling, hypoalgesia or analgesia).
- Age: 18 to 75 years.
- Consent to participate in the study.

Similarly, the exclusion criteria established are as follows:

1. Previous surgery on the wrist
2. Acquired or hereditary pathology causing peripheral neuropathies
3. Polyneuropathy of any etiology
4. Rheumatoid arthritis
5. Diabetes Mellitus

## Interventions

### Intervention description {11a}

We work side by side with the neurophysiology service of the Virgen Macarena hospital (Seville, Spain); they will apply the neurophysiological evaluation protocol between 1 and 3 months previous to the echography study.

#### Neurophysiological

The neurophysiological evaluation protocol is based on the recommendations of the AAEM (12), listed in table 1.

**Table 1.** Recommendations of the American Association of Electrodiagnostic Medicine (AAEM)

| PROTOCOL FOR NEUROPHYSIOLOGICAL STUDY ACCORDING TO AAEM <sup>(12)</sup> |  |
| --- | --- |
| **Parameter 1** | Performing median motor conduction velocity with surface electrodes (recording on abductor pollicis brevis-APB- and reference along the distal tendon-extremus thumb). Bipolar electrode stimulation at wrist and elbow. Calculation of distal motor latency (DML), motor conduction velocity (MCV) wrist-elbow. Assessment of amplitude and duration of the potentials obtained. |
| **Parameter 2** | Ulnar motor conduction velocity with surface electrodes (recording in the hypothenar eminence and reference along the distal tendon-extremus-fifth finger). Bipolar electrode stimulation at wrist and elbow (inferior-superior). Calculation of distal motor latency, motor conduction velocity wrist-lower elbow and lower elbow-upper elbow. Assessment of amplitude and duration of the potentials obtained. |
| **Parameter 3** | Orthodromic median sensory conduction with stimulation on 2 and 4 fingers and palm and recording on wrist. Stimulation on 3 fingers and recording on palm. Assessment of sensory conduction velocity (SCV) in the 2 finger-wrist, 4 finger-wrist, palm-wrist and palm-wrist segments. Assessment of amplitude and duration of the potentials obtained. |
| **Parameter 4** | Ulnar orthodromic sensory conduction with stimulation on 4 and 5 fingers and recording at the wrist. Assessment of conduction velocity in the 4-wrist |
|  | and 5-wrist segments and amplitude and duration of the potentials obtained.<br>Comparison of the IV middle finger-ulnar. |
| **Parameter 5** | Orthodromic radial sensory conduction with stimulation of the dorsum of the MCF joint of 1 finger and recording on the radial edge of the forearm.<br>Assessment of conduction velocity and amplitude and duration of the potentials obtained. |
| **Parameter 6** | Antidromic sensory conduction of median, ulnar and radial with surface electrode recording at the base of 2 and 4 fingers (median), at the base of 4 and 5 fingers (ulnar) and at the dorsum of 1 finger MCF (radial) with stimulation at the wrist for median and ulnar, and at the radial edge for radial. |

#### Ultrasound

In our study we followed the ultrasound protocol described by Chen et al. (11) (table 2), discarding dynamic parameters and adding others based on the most recent literature.

**Table 2.** Ultrasound protocol

| **VARIABLES** | **DESCRIPTION** |
| --- | --- |
| General Position | The patient is seated facing the examiner with the forearm resting supinated on a stable surface (for other patients it may be more comfortable to lie face down with the forearm in full pronation).<br>A rolled towel is placed under the wrist to allow 30°-45° extension of the wrist, minimizing the thenar eminence for better transducer glide. The use of a generous amount of gel is helpful. (*) |
| Nerve cross-sectional area (CSA) in distal forearm | In the transverse axis, in the distal and volar third of the forearm, the pronator quadratus is located between the radius and ulna. The CSA for the calculation of the wrist-forearm index is measured at this level. (**) |
| CSA at tunnel entrance | At the level of the pisiform the cross-sectional area of the nerve is measured. |
| CSA at the tunnel exit and mass search | The cross-sectional area of the nerve is measured at the level of the hook of the hamate.<br>At the same time, an anatomical analysis is carried out to look for any space-occupying lesions (accessory lumbricals, tenosynovitis or hidden cysts, etc.). |
| Longitudinal axis of the nerve at the exit of the tunnel | Rotate the transducer in line with the 3rd metacarpal; nerve compression may show as a notch or change in caliber. The carpal tunnel is also examined for space-occupying masses by short-axis sliding along the full width of the tunnel. |
| Dynamic test 1: Thenar digital flexion stress test | Compressing a soft object (towel) between the 1st and 2nd finger is visualized transversely and longitudinally; positive result due to flattening of the nerve due to compression of the flexor tendon. No CSA measurement. |
| Dynamic test 2: Superficial digital flexor digitorum intrusion test | The superficial and deep flexor belly should enter the tunnel with the wrist in hyperextension contributing to nerve compression. No CSA measurement. |
| Dynamic test 3: Intrusion test lumbrical muscles | To be positive, flexion of the fingers with full grip must enter the tunnel at the muscle belly of the lumbricals compressing the nerve. No CSA measurement. |

(\*) in our case, the cylindrical gel canister is the one we have positioned under the patient's wrist to achieve its hyperextension.
(\*\*) it should be noted that for the measurement of the wrist-forearm index we have carried out, based on other publications, the HAG at the entrance of the tunnel between the HAG and 12 cm from the distal crease of the wrist.

### Criteria for discontinuing or modifying allocated interventions {11b}

Not applicable since the intervention comprises a one-time diagnostic procedure.

### Strategies to improve adherence to interventions {11c}

Not applicable since the intervention comprises diagnostic procedure.

### Relevant concomitant care permitted or prohibited during the trial {11d}

No concomitant care was prohibited during the study.

### Outcomes {12}

1. **Demographic variables**: sex, age, time of evolution of the clinic, manual dominance, hand affected, body mass index, employment status, type of work (pink, white or blue collar), time worked, triggers of symptoms, what symptoms are relieved with, as well as toxic habits, whether they are smokers (to be answered affirmative or negative) or alcohol consumers (to be answered affirmative or negative).
2. **Electrodiagnostic variables**: motor conduction velocity of the median nerve (calculating distal motor latency and wrist-elbow motor conduction velocity), motor conduction velocity of the ulnar nerve (calculating distal motor latency, lower and upper wrist-elbow motor conduction velocity), orthodromic sensory conduction of the median, ulnar and radial nerve, as well as antidromic sensory conduction of the median, ulnar and radial nerve.
3. **Ultrasound variables**:

- The cross-sectional area (CSA), to be performed with linear tracing, measured over the inner border of the epineurium of the median nerve at the level of the entrance and exit of the carpal tunnel. We take the average of 3 consecutive measurements.
- The range of median nerve thinning understood as the ratio of the transverse and anteroposterior diameters of the nerve in the proximal-medial intracarpal region, at the level of the pisiform.
- The wrist-forearm index, understood as the ratio of the CSA at the tunnel entrance to the CSA at 12 cm from the distal wrist crease.
- The flexor retinaculum bulge, which translates an increase in intracarpal pressure. We measure this by tracing a horizontal from the superior pole of the scaphoid to the superior pole of the pisiform, and a vertical from the superior end of the flexor retinaculum to the horizontal.
- Power Doppler uptake, which determines the hypervascularization of the nerve, especially in the
- periphery, at the expense of low-flow intraneural vessels. The uptake is at mid-tunnel level, between the pisiform and hamate nerves.
- The existence of notches in the nerve in the longitudinal ultrasound view, including the entrance, interior and exit of the tunnel.
4. **Boston Carpal Tunnel Questionnaire** (BCTQ), to assess both the symptoms and the subjective functional impact of patients.
5. **Cost evaluation**: To carry out a cost-effectiveness analysis, we will evaluate the costs of the consumables established by the Andalusian Health Service used in both tests and the average time for each of the tests, in order to calculate the cost of the professional when performing each of them.

### Participant timeline {13}

As shown in table 3, we have established a first period for the study, from July 2020 to December 2020, where the initial literature search is carried out, the study protocol is drawn up and submitted to the ethics committee for approval. During this time, a training period is also planned for the researcher in the performance of the ultrasound protocol, a study carried out in the Physical Medicine and Rehabilitation Services of the Virgen Macarena, Virgen del Rocio and Valme University Hospitals, where ultrasound studies are performed on patients with CTS.

**Table 3.** Participant timeline

| Staff responsible | Task | First period | Second period | Third period |
| --- | --- | --- | --- | --- |

|  |  | From July 2020 to<br>December 2020 | From January 2021<br>to September 2022 | From September<br>2022 to May 2023 |
| --- | --- | --- | --- | --- |
| Principal investigator<br>and sponsors | Bibliographic search | X |  |  |
| Principal investigator<br>and sponsors | Protocol development | X |  |  |
| Principal investigator<br>and sponsors | Ethics Committee | X |  |  |
| Principal investigator<br>and sponsors | Training period | X |  |  |
| Principal investigator | Ultrasound study<br>performed on the<br>sample |  | X |  |
| Principal investigator | Data collection |  |  | X |
|  | Statistical analysis |  |  | X |
| Principal investigator | Writing |  |  | X |
| Sponsors | Corrections |  |  | X |

During the second period of the study, from January 2021 to December 2021, we planned to have performed ultrasound studies on patients. However, due to the Covid-19 pandemic situation, we were forced to prolong this period until September 2022.

We propose a third and final period from September 2022 to May 2023 for data collection, statistical analysis and writing of the study, as well as the relevant corrections.

### Sample size {14}

To calculate the sample size (EPIDAT program) we proposed:

- Expected sensibility: 78%
- Expected specificity: 87%
- Level of confidence: 95%

**Table 1.**
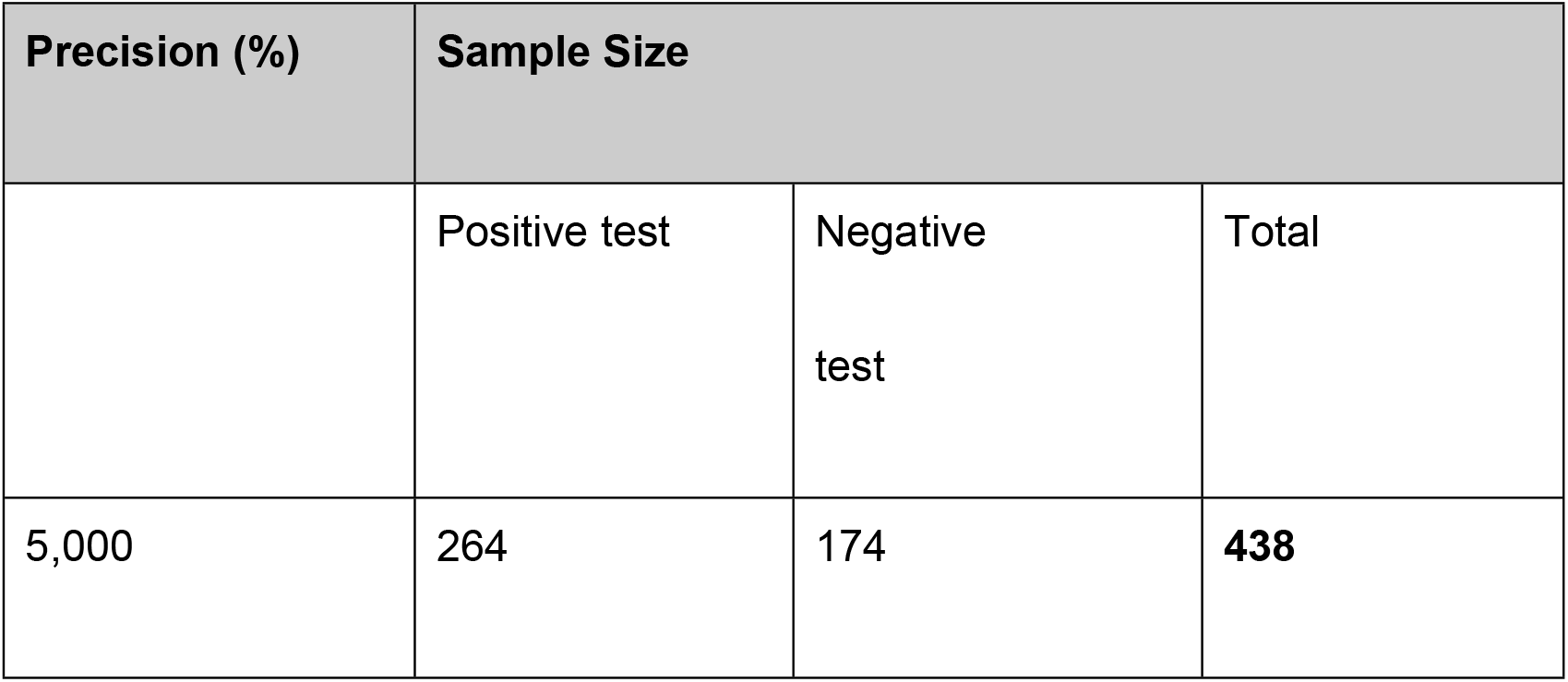
Sample size

In order to evaluate ultrasound as a diagnostic test versus neurophysiological study in patients with a high clinical suspicion of suffering from CTS, expected values of sensitivity and specificity of the ultrasound test of 78% and 87% respectively, a maximum imprecision of 5% and a confidence level of 95% are considered, resulting in the need to evaluate a minimum of 264 patients with a positive result in the nerve conduction study as well as a minimum of 174 patients with a negative result, which means a total sample of 438 patients. The calculation was performed using the EPIDAT program.

### Recruitment {15}

The patients included in the study will be obtained consecutively through the consultations of the Neurophysiology Department of the Virgen Macarena Hospital (Seville, Spain), provided that they have clinical suspicion of CTS.

## Methods: Assignment of interventions (for controlled trials)

### Sequence generation {16a}

Not applicable, since there are no intervention groups. All patients we recruited from the neurophysiology department of the Vigen Macarena Hospital (Seville, Spain), who underwent an ultrasonography following a specific protocol.

### Concealment mechanism {16b}

Not applicable, since there are no intervention groups.

### Implementation {16c}

Not applicable, since there are no intervention groups.

## Assignment of interventions: Blinding

### Who will be blinded {17a}

Not applicable. This protocol is for an evaluation study of diagnostic tests comparing electroneurography and ultrasound. Nevertheless, the result of the neurophysiological study will be blinded so the researcher does not know the expected outcome of the ultrasound.

### Procedure for unblinding if needed {17b}

Not applicable. No unblinded procedure will be carried out due to the diagnostic nature of the study.

## Methods: Data collection, management, and analysis

### Data collection methods

#### Plans for assessment and collection of outcomes {18a}

The means necessary to carry out our study will be a Sonosite ultrasound scanner model X-porte, ultrasound gel and a computer for data collection in Excel table. We do not need external funding to carry out the study.

In order to promote data quality, a training period was included for the researcher in charge of performing the ultrasounds, it was carried out during the first period of the timeline.

On the other hand, when the ultrasound was performed, 3 measurements were made of each parameter, taking the average of the three of them.

Of the instruments included in the study, the following should be highlighted:

- Boston Carpal Tunnel Questionnaire – (BCTQ): The Boston scale is a validated and self-administered instrument that assesses the impact of CTS on both symptoms and patient function (13), having demonstrated a statistically significant relationship between AST, NF, clinical symptoms and hand functionality (14,15). Contrary to the above, Mondelli et al. in 2008 (16) obtained results when comparing the combination of ultrasound and NF versus BCTQ, finding no relationship between the questionnaire and the other two tools. However, more publications support the usefulness of BCTQ, being a tool translated into several languages such as Arabic, Turkish or Japanese (17). Of the tests used in the physical examination the following should be highlight:
  - Tinel sign: The sensitivity is between 26 and 79% and the specificity is between 40 and 100% (18).
  - Phalen sign: the sensitivity is between 67 and 83% and the specificity is between 47 and 100% (18,19).

### Plans to promote participant retention and complete follow-up {18b}

Not applicable. The data collection is a one-time ultrasound and the are no further revisions or assessments scheduled after.

### Data management {19}

The data will be entered by one of the two principal investigators, in batches of 50 patients, performing a subsequent review of the data to ensure that there are no errors when entering them.

We will not use the patient’s personal data but the personal health code (NUSHA).

The storage will be done in Excel format on a computer and on an external storage system (USB).

## Statistical methods

### Statistical methods for primary and secondary outcomes {20a}

After an initial exploration of the data, quantitative variables are expressed as means and standard deviations or medians and quartiles in case of asymmetric distributions, and qualitative variables as percentages. The calculation of the measures of validity and safety of the ultrasound test as a diagnostic test for CTS: sensitivity, specificity, predictive values and likelihood ratios, are carried out considering the neurophysiological study as the reference test (gold standard).

Likewise, the reliability of the ultrasound is analyzed by calculating the degree of agreement between the diagnoses of the two tests using Cohen’s Kappa coefficient. The concordance of both tests with the established clinical severity is also studied.

The analysis of the ultrasound parameters predictive of CTS is approached univariate with the Student t-test for two independent samples or Mann-Whitney U test for quantitative variables and with the chi-square test or non-asymptotic methods for qualitative variables.

Shortly afterwards, a multivariate logistic regression model will be created to try to determine the best subset of them that predict CTS.

Finally, ROC curves will be performed to evaluate cut-off points for certain quantitative ultrasound parameters that discriminate well against CTS.

Data analysis is performed with SPSS 25.0 statistical software.

### Methods for additional analyses (e.g. subgroup analyses) {20b}

Not applicable, no additional analysis will be performed.

### Methods in analysis to handle protocol non-adherence and any statistical methods to handle missing data {20c}

The main analysis will be the concordance between the results of electroneurography and ultrasound in the diagnosis of CTS. Different parameters will be considered and in case of missing values these will be imputed by multiple imputation, selecting some of the methods that best suits the data set, these being the Monte Carlo method and Markov chains (MCMC), the maximum interaction method, monotonic method, linear regression, predictive mean matching (PMC) among others.

## Methods: Monitoring

### Data monitoring

#### Composition of the data monitoring committee, its role and reporting structure {21a}

We did not need a data monitoring committee as a single measurement of the variables will be included in the study, and the serial collection of the variables was not necessary. Each patient was explored, the questionnaire was passed to them, and neurophysiology and ultrasound were performed on a single occasion.

#### Interim analyses {21b}

No interim analysis will be performed.

### Harms {22}

Although the diagnostic tests evaluated in this study are harmless for humans, we have to consider the possibility or adverse effects, and how we will collect them when applying the test throughout the study. On the one hand, at the end of the ultrasound, we will collect the appearance of any adverse effect by answering a Yes / No question, and if so, a brief description of it. If the adverse effect come up hours after the test was carried out, all patients have, in their informed consent copy, an email address of the principal investigator that they can attend.

Regarding the possible side effects derived from the neurophysiological test, being a different department from Rehabilitation, Regarding the possible side effects derived from the neurophysiological test, being a different department from Rehabilitation, it is they who manage the appearance of these adverse effects as part of their daily clinical activity.

### Auditing {23}

Of the two main researchers, one of them will carry out the ultrasounds and data collection, while the other one will carry out the quality control of the study, a well-done data collection, the proper functioning of the ultrasound system, the appointment of patients, etc.

## Discussion

Ultrasonography allows not only to objectify median nerve alterations but also the underlying pathological mechanisms in CTS. The median nerve undergoes physiological changes including swelling and edema as it is compressed within the tunnel, resulting in increased CSA and hypoechogenicity. As the nerve becomes inflamed, it moves against the flexor retinaculum which becomes bowed, further compressing the nerve. In advanced CTS, inflammation of the nerve leads to hypervascularisation, which can be detected by Doppler. In turn, ultrasound can also be useful to assess underlying structural causes of CTS, such as cysts, ganglions or the presence of other masses invading the tunnel (20). These structural changes that the nerve undergoes are translated into ultrasound parameters that we can measure, and which position ultrasound as a useful tool in the study of the syndrome. It is important to note that in early stages, morphology may not be affected, so in the presence of normal ultrasound findings in patients with suggestive symptoms, we should not exclude the diagnosis of CTS (21).

Regarding the sensitivity and specificity of ultrasound, in 2009 Beekman and Viser observed a sensitivity of 70-88% and a specificity of 57-100%. In turn, in the meta-analysis carried out by Fowler et al. (22), although with a debatable methodology, they found similar results, with a combined sensitivity of 80.2% and a specificity of 78.7% (using neurophysiological testing as a reference). Others give sensitivity figures of 78% and specificity of 87% (23), with values of up to 94% sensitivity and 98% specificity having been described (24).

**Figure 1.**
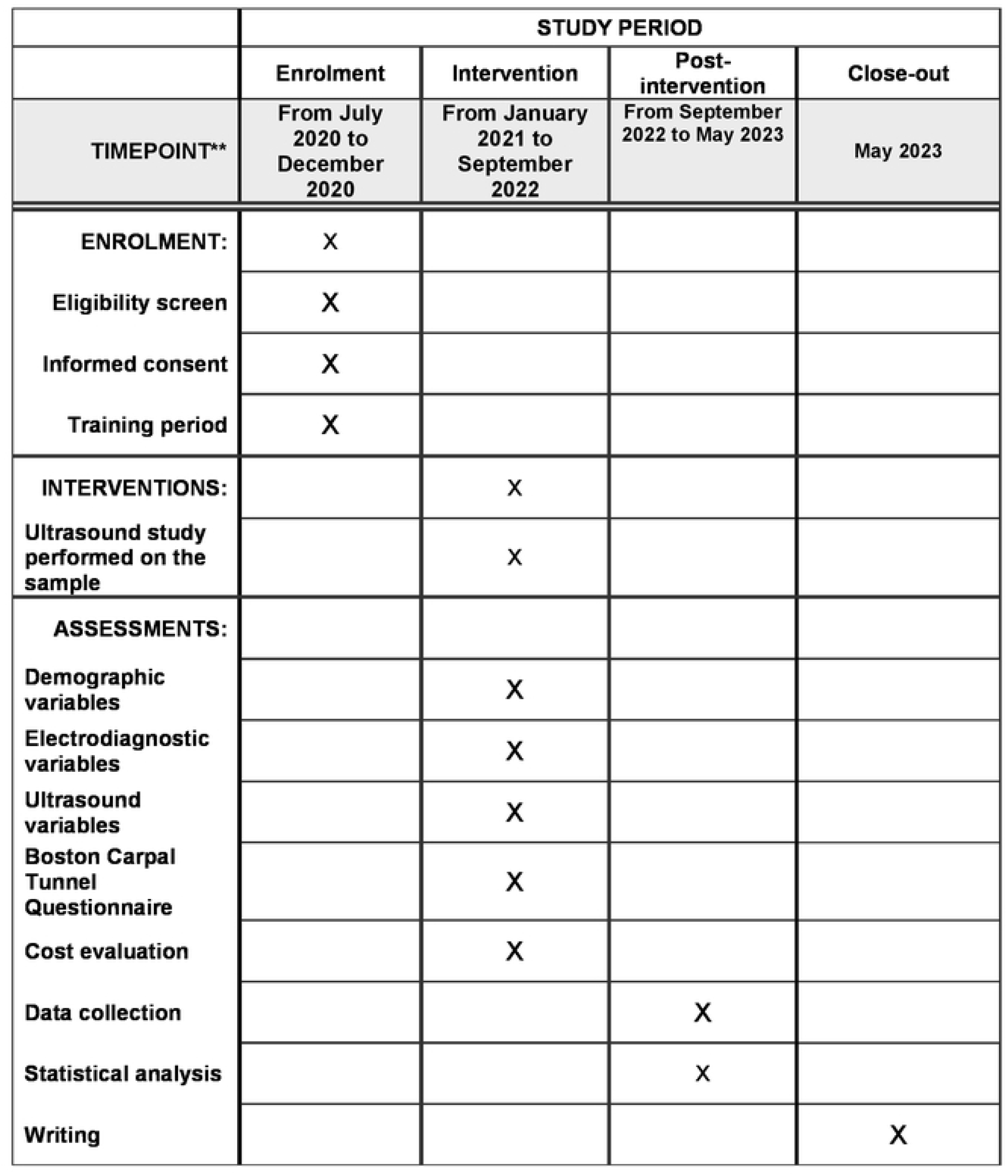
Schedule of enrolment, interventions, and assessments.

It should be emphasized that ultrasound is operator dependent (26), therefore good training is very important to ensure reliability and reproducibility.

If we analyze the ultrasound assessment protocols described to date, there has been much disparity between scientific publications; like Roll et al. (25), we consider this to be the main reason why there is no standardized protocol for ultrasound study as a diagnostic test.

The authors Chen et al. (11) proposed an ultrasound assessment protocol that included:

- Patient position.
- Ultrasound parameters: CSA at the entrance and exit of the tunnel, CSA in the distal third of the forearm, longitudinal view of the nerve, and dynamic tests (when flexing the first finger and when flexing the rest of the fingers) both to observe changes in the diameter of the nerve without measurement, and lumbrical tests (to determine whether they invade the tunnel when making a full grip).

In our study, we have based ourselves on the general lines described in this protocol, adding other parameters based on the literature review.

Among the ultrasound parameters analyzed, Duncan et al. were the first to demonstrate the superiority of the CSA over the other ultrasound parameters (26), although there are authors (27) who have suggested the superiority of the nerve inflammation ratio over the rest. In turn, Swen et al. established that the optimal atomic reference for measuring CSA is the entrance of the tunnel at the level of the pisiform (28).

As Hobson-Webb et al. (29) have already stated, a single measurement can result in a false positive, so in our protocol we have decided to take the mean of 3 consecutive CSA measurements of the median nerve, but not for the indices or retinaculum ballooning.

Regarding the cut-off point for CSA, a lot of variability is reported in the literature. CSA values have been reported from as low as 6.5 mm2 (30) to as high as 13 mm^2^ (31), with sensitivity ranging from 60% (32) to 100% (33) and specificity ranging from 22% to 100%, as well as overall accuracy ranging from 68% (28) to 97.2% (34).

Considered normal limits of nerve CSA range from 7 to 9.4 mm2 (30), and diagnostic values vary from 9 to 15 mm2 (36). This variability is mainly due to the different measurement techniques used between studies. It does seem clear that the area of the median nerve increases as the syndrome progresses over time (37).

Descatha et al. (38) conducted a meta-analysis of 13 articles in 2012 and observed that for a cut-off point between 7-8.5 mm^2^ the sensitivity amounted to 94% (0.94 [0.87-1.00]) with a likelihood ratio for the negative test of 0. 15 [0.08-0.30]; similarly, for a cut-off point between 11.5-13.00 mm^2^ the specificity amounted to 97% (0.97 [0.91-1.00]), with a likelihood ratio for the positive test of 8.5 [2.83-25.57]. On this basis, they state that ultrasonography cannot be considered as an alternative to the neurophysiological study, but it can be used as a screening tool.

Similarly, and in relation to the diagnostic accuracy of the CSA parameter, the most extensive review carried out in this regard dates from 2012 by Cartwright et al. (39), who reviewed 121 articles and concluded that ultrasound “probably adds value to nerve conduction studies in the diagnosis of CTS for detecting associated anatomical alterations” and recommended “using ultrasound as a screening tool to assess structural alterations in patients with suspected CTS”.

In our study, we included both patients with CTS (confirmed by neurophysiological testing) and patients considered “healthy” (with negative neurophysiological testing), in order to analyze the ultrasound differences between the two groups.

Hobson-Webb et al. (29) observed that the CSA in healthy patients was lower than in patients with CTS, with values of 10 mm2, higher than that reported by authors such as Hammer et al. (35) or Yesildag (40), but similar to that reported by others such as Nakamichi and Tachibana (41). Similarly, Hobson-Webb et al. also observed that AST in the forearm differed between healthy (9.8 ± 2.4 mm^2^) and diseased (6.9 ± 1.6mm^2^).

In relation to the wrist-forearm index, the first to consider this parameter were Hobson-Webb and his team, who observed that in normal subjects the CSA in the wrist has been described as having the same value as that of the forearm and hypothesized that the ratio between the two could be a diagnostic alternative (29), on this basis, the wrist-forearm index should be 1:1.

Both Hobson-Webb et al. (29) and Mhoon et al. (42) observed that values of 1.4 or higher have a high sensitivity (97-100%) to diagnose CTS and values below 1.4 a high sensitivity (99%) to rule it out, the low specificity (28%) limiting its use to screening only.

In our study, based on the approach of Hobson-Web et al. (29), we measured CSA at the tunnel entrance and 12 cm from the distal wrist crease.

There are authors who have performed this index using different measurement references (43), so we will not, at the time, establish a comparison between their findings and those of our study. We took 1.4 as the cut-off point, as it is the most frequently considered cut-off point.

A case-control study conducted by Fu et al. in 2015 (44) performed the entrance-exit index (tunnel entrance CSA /exit CSA) in patients with clinically suggestive and positive electroneurogram, observing that for index values greater than or equal to 1.3, a sensitivity of 93% and specificity of 91% is obtained to diagnose CTS. Like us, they consider that the index can compensate for interexplorer variability when measuring CSA; they also observed that the index is not related to demographic variables and can be used without a population condition. In our study we considered a cut-off point of 1.3.

Echogenicity is another ultrasound parameter that has been studied in possible relation to the development of CTS, with a decrease in echogenicity observed as the syndrome progresses (45). Under normal conditions, a healthy median nerve is observed in transverse plane with high echogenicity, with its fascicles translating into the so-called “honeycomb” pattern, which is altered especially at the level of compression (46). However, this parameter is very subjective even in experienced examiners, and may vary depending on the ultrasound scanner used, so we have dispensed with this parameter in our study.

In relation to the Doppler tool, its usefulness lies in the fact that in healthy subjects intraneural blood flow has never been demonstrated, which seems to reflect a pathological condition. It has been observed that Doppler can detect increased intraneural or epineural vascular flow in patients with CTS, which seems to reinforce the diagnosis (47), and a correlation has been observed between intraneural flow and CTS severity (48).

There are 3 Doppler techniques capable of detecting blood flow: color, power and spectral Doppler. Of these, power doppler is not affected by compression or alising when detecting blood flow, since blood volume (rather than velocity) is what is measured. In addition, power doppler is more sensitive when flow is slow (49).

When applying the doppler, the position of the wrist is very important, which could also influence blood flow; the authors Vanderschueren et al. recommend keeping the wrist in a neutral position, avoiding mobilization of the fingers or provocation tests (50).

Mohammadi et al. reported that the presence of color Doppler uptake in the median nerve correlates strongly with the severity observed in neurophysiological findings (48).

Roll et al. (25) developed a model with severity stages based on doppler uptake in patients with CTS; an 8-point system (from 0 to 7) taking the neurophysiological outcome as the gold standard. They mention that the assessment is more accurate in moderate stages.

Superb microvascular imaging (51), which is a pioneering technology in ultrasound that allows real-time blood flow assessment, developed by Toshiba, has also been described as far superior to the Doppler tool. However, it is not currently accessible to physicians, and therefore cannot be taken into consideration.

Another parameter studied over time has been the thickening of the flexor retinaculum in the proximal region of the carpal tunnel, the interest being based on the fact that continued inflammation and the consequent extraneural reactive fibrosis cause a greater thickness of this structure, considered in the pathological range above 1 mm (52). In any case, in our opinion, like Pardal-Fernández (53), we consider it to be a parameter of low sensitivity, due to the small size of its dimensions and the consequent variability in measurements.

The flattening ratio is a ratio between the transverse and anteroposterior diameters in the intracarpal region, described within the tunnel. It is a well-regarded parameter (54) and ratios above 3.5-4 are considered pathological. At the mid-distal level, in the ulnar nerve, some authors have shown that the trapped nerve presents a morphological deformation of its floor, in such a way that it outlines the contour or ridge of the tendons it contacts, showing a deformity similar to a “triangular” (55), which is produced by the lower physical resistance of the nerve to the impaction of the tendons, which are more rigid and consistent. In turn, the authors Lee et al. carried out a prospective case-control study analyzing different ultrasound variables among which they included the flattening index, but at different levels of the tunnel: at the pisiform, hamate and lunate; they only obtained a statistically significant difference between both groups in the measurement made at the level of the hamate (cut-off point 3. 2+/-0.4 P<0.05) (56); these same results were found by Tai et al. in a meta-analysis carried out in 2012 (57). In any case, some authors did not find cost-effectiveness in this parameter (58). We have included it in our parameters and hope to clarify its usefulness.

The flexor retinaculum may bulge as a result of increased intracarpal pressure. The buckling of the flexor retinaculum, in most CTSs, is usually not important considering the low elasticity of this structure, especially if it is also fibrosed. It is observed more in the distal third, at the level of the hamate bone, and is established by determining the distance from the crossing of the line that joins the upper edge of the trapezium bone with that of the hamate bone, with the line that goes from the floor, the large bone, to the highest point of the vault of the retinaculum or anteroposterior diameter. It is considered pathological if it is greater than 2 mm (52), although some consider it to be 3.7 mm (55). In any case and taking into account that the image at this point is not usually good, these findings are not very conclusive due to their low reproducibility. We have included this parameter in our measurements, finding it useful based on the literature analyzed and hoping to clarify the best cut-off point.

Another of the ultrasound parameters described in the literature is the overall dimension of the tunnel (43), understood as the ratio between the scaphoid-pisiform distance and the distance from the flexor retinaculum to the lunate; a more square morphology has been described in patients with CTS compared to healthy patients (43). This parameter, although included in the literature, has not been studied or taken into account as much as others, so we have not included it in our study in order to propose a protocol that is as useful and simple as possible for its application in clinical practice.

The mobility patterns of the nerve in patients with CTS have also been analyzed. The authors Kuo et al. (59) studied the transverse gliding patterns of the median nerve during finger movement, analyzing dynamic images to distinguish between healthy subjects and affected patients; they also proposed a formula to be integrated into the ultrasound software for assessment.

Chen et al. (11) also included in their protocol a “stress test” when flexing the first finger and when flexing the rest of the fingers (changes in the diameter of the nerve without measuring) as well as the lumbrical test (to determine whether they invade the tunnel when performing a full grip). We found the performance and assessment of these tests to be highly dependent on the experience of the examiner, which is why we have dispensed with dynamic tests in the development of our protocol.

The last two “items” that we analyzed in our study are, on the one hand, the search for anatomical alterations that may compress the median nerve as it passes through the tunnel (ganglions, tendonitis, etc.), since ultrasound allows easy detection of anatomical alterations of the nerve (41). In our study, as described above, we performed a scan of the image in both the transverse and longitudinal planes, covering all structures from lateral to medial.

On the other hand, we assessed the presence of nerve grimacing in the longitudinal plane, to identify whether there is a specific point of compression. In general, the nerve increases in size immediately after the compression zone.

Other variables included in our study are demographic variables; it is in our interest to analyze whether there is a predisposition to suffer from CTS in certain subjects according to their baseline characteristics. This has been studied previously (60, 61), and it has been observed that weight is directly related to the presence of the syndrome.

We have included the BCTQ scale, a validated, self-administered instrument that assesses the impact of CTS on both symptoms and patient function (62), and a statistically significant relationship has been demonstrated between CSA, NPT score, clinical symptoms and hand function (14, 15). Contrary to this, Mondelli et al. in 2008 (16) found no relationship when comparing the combination of ultrasound and NPT score versus BCTQ.

However, there are more publications that support the usefulness of the BCTQ, being a tool translated into several languages such as Arabic, Turkish or Japanese (17).

When comparing neurophysiological studies versus ultrasound, although the former offer 85% sensitivity and 95% specificity in the diagnosis of CTS, they are invasive and unpleasant for the patient (12); recent literature has shown that patients themselves have a preference for ultrasound over nerve conduction studies (62).

Although the specificity of neurophysiological tests is high, there is significant variability in their sensitivity (56-85%) which can translate into false negative results between 10-20% of patients (39) and false positives between 16-34% (22). Other authors report an NPT sensitivity of 88% and specificity of 93% (54).

Seror in his 2008 review concluded that there is no “competition” between ultrasound and NPTs, but rather a complementarity between the two tests (63).

Although ultrasound may not replace neurophysiological testing as the gold standard for confirmation, authors Fowler and Gaughan suggest that it could be proposed as a first-line tool (22); they add that the best candidates for ultrasound are those with high prior probability based on examination and reported symptomatology.

Several authors (38,39) propose ultrasound as an initial screening test, leaving nerve conduction for definitive diagnosis.

The use of ultrasound as a diagnostic tool in CTS has many advantages for both the physician and the patient. It is a non-invasive, convenient, and fast tool, and ultrasound equipment is becoming increasingly cheaper, making it more accessible to the healthcare system (20).

## Data Availability

No datasets were generated or analysed during the current study. All relevant data from this study will be made available upon study completion

https://clinicaltrials.gov/ct2/show/NCT05556278

## Ethics and dissemination

### Research ethics approval {24}

This study has been approved by the Andalusian Biomedical Research Ethics Committee (Comité de Ética de la Investigación Biomédica de Andalucía). Written informed consent will be obtained from all participants before entering in the study.

### Protocol amendments {25}

We will communicate it by email with the relevant parts, as the secondary investigators or the statistician.

### Consent or assent {26a}

The patients included in the study will be obtained consecutively through the consultations of the Neurophysiology Department of the Virgen Macarena Hospital (Seville), provided that they have clinical suspicion of CTS. These patients will be invited to participate in the study and will be asked to sign the informed consent form, which reflects the documentary commitment to confidentiality and safeguards in the treatment of their data by the researchers, as well as information on what their participation in the study will consist of.

### Additional consent provisions for collection and use of participant data and biological specimens {26b}

Not applicable, as these will not be collected.

### Confidentiality {27}

In order to protect participants personal information, we did not use their names, but their personal health code called “NUSHA” (e.g.: AN0123456789).

### Access to data {29}

Only the two main investigators will have access to the final trial dataset as well as the statistician who will handles the database.

### Ancillary and post-trial care {30}

Being the echography an innocuous test, we do not stand any provision for post-trial test. Not applicable.

## Dissemination policy

### Dissemination plans {31a}

The trial results will be communicated to the scientific community through journal as well as communications at scientific meetings or congresses. As for the patients participating in the trial, again within the informed consent they will have an email address to contact with in case they want to know the results; this way we do not bother those participants who do not want to know the results.

### Authors’ contributions {31b}

MPMC is the principal investigator, having developed the idea and protocol with the help of the research team. She will be responsible for collecting the sample. MRPD led the study proposal and developed the protocol, being the main methodologist. ASJS participated in the development of the protocol until the final version was reached. MAL facilitated the collection of the sample by performing the neurophysiological tests in the first stage, and by bringing the patients to the principal investigator. In turn, she provided the neurophysiological protocol used to be included in the manuscript. GJJ facilitated the collection of the sample by performing the neurophysiological tests at an early stage, getting the patients to the principal investigator.

### Plans to give access to the full protocol, participant level-data and statistical code {31c}

The full study protocol can be accessed by contacting the trial team at (e-mail). Anonymized participants level data and statistical codes used for the analysis are available on request and subject to data sharing agreements. Data sharing may only be possible after relevant embedded trials have been published.

## Trial status

This protocol is version 4 after several corrections and modifications based on the most recently published literature. We will start recruitment in January 2021 and expect to complete it in 2023.

## Abbreviations

CSA: Cross sectional area.
AAEM: American Association of Electrodiagnostic Medicine.
BCTQ: Boston Carpal Tunnel Questionnaire
CTS: Carpal Tunnel Syndrome.
NPT: Neurophysiological test.

## Acknowledgements

We thank Merz Pharma for assisting in the development of the translation of the manuscript and appendices.

